# Enhancing public health syphilis screening program: a comparative analysis between four different immunoassays

**DOI:** 10.1101/2025.03.16.25324064

**Authors:** Gheyath Nasrallah, Nadin Younes, Jawaher A. Al-Emadi, Hadiya M. Khalid, Manal Elshaikh, Mazen Najib Abouassali, Ibrahim Wissam Karimeh, Mohammed Abdelfatah Ibrahim, Mutaz Mohamed Ali, Ibrahim Al Shaar, Parveen Banu Nizamuddin, Salma Younes, Hadi M Yassine, Laith J. Abu-Raddad, Ahmed Ismail

**Affiliations:** Department of Biomedical Science, College of Health Sciences, QU Health, Qatar University, 2713 Doha, Qatar; Biomedical Research Center, Qatar University, 2713 Doha, Qatar; Laboratory Section, Medical Commission Department, Ministry of Public Health, Doha, Qatar; Infectious Disease Epidemiology Group, Weill Cornell Medicine-Qatar, Cornell University, Qatar Foundation—Education City, 24144 Doha, Qatar; World Health Organization Collaborating Centre for Disease Epidemiology Analytics on HIV/AIDS, Sexually Transmitted Infections, and Viral Hepatitis, Weill Cornell Medicine-Qatar, Cornell University, Qatar Foundation-Education City, 24144 Doha, Qatar; Department of Healthcare Policy and Research, Weill Cornell Medicine, Cornell University, New York, NY 10065, USA

**Author notes:** Correspond to: Ahmed Ismail, Consultant Clinical Pathologist and Section Head Laboratory, Medical Commission Dept, Ministry of Public Health, Doha, Qatar, Gheyath K. Nasrallah, associate professor, Department of Biomedical Science, College of Health Sciences, Qatar University, Doha, Qatar.

**Keywords:** Treponemal antibodies, Syphilis, CLIA, Snibe, Abbott

## Abstract

**Background:** Syphilis remains a significant global public health concern, contributing substantially to morbidity and mortality. Given syphilis’ asymptomatic nature, varied clinical presentations, and severe untreated complications, accurate diagnostic methods are crucial.

**Aim:** This study aims to evaluate the performance of the new treponemal test, MAGLUMI-X3® Ab-syphilis assay (Maglumi-TP) from Snibe Diagnostics, China, as a potential sensitive, specific TP-Ab-screening & confirmatory method by comparing its performance to the (i) well-established and FDA-approved Abbott ARCHITECT Syphilis-TP (Architect-TP) assay and (ii) Fujiribio INNO-LIA® Syphilis Score (INNO-LIA-TP) line-immunoassay. In addition, to evaluate the false positive rate of the RPR non-treponemal test. In this study, we proposed Architect-TP and INNO-LIA-TP as reference assays.

**Method:** We selected 180 samples that demonstrated agreement and discrepancies between the RPR and Architect-TP. This selection comprised 40 cases (RPR-TP + Architect-TP +), 40 cases (RPR-TP + Architect-TP -), and 100 cases (RPR-TP – Architect-TP -). All selected samples underwent re-testing using the Maglumi-TP and the INNO-LIA-TP assays.

**Results:** Maglumi-TP demonstrated excellent sensitivity and specificity of 100%, with perfect agreement (*κ*= 1.00) with Architect-TP. Similarly, Maglumi-TP exhibited excellent sensitivity of 100% (95% CI: 91-100), specificity of 100% (95% CI: 97.4-100), and perfect agreement (*κ*= 1.00) with the INNO-LIA-TP. Notably, 40 cases (28.8%; 40/139) exhibited false-positive results when using the RPR test compared to the INNO-LIA-TP, indicating a substantial false-positive rate associated with the RPR assay.

**Conclusion:** With its high sensitivity and absence of false-positive and false-negative results, Maglumi-TP emerges as a promising automated screening assay for detecting and confirming TP antibodies.

## 1. Introduction

Sexually transmitted diseases (STDs) pose a persistent global public health challenge, significantly contributing to morbidity and mortality, particularly in resource-limited settings (1). Among these diseases, syphilis, caused by the bacterium *Treponema pallidum* (*T. pallidum; TP*), stands out as one of the oldest and most enduring infections, exerting substantial impacts on global quality of life, health, and economies (2). The natural course of syphilis encompasses primary, secondary, latent, and tertiary stages, with the primary, secondary stages, and early latency being the most infectious (3). If left untreated, syphilis can lead to severe complications, causing permanent damage to the nervous and cardiovascular systems and often resulting in life-threatening consequences (4). Annually, an estimated 6 million new cases of syphilis are reported globally in individuals aged 15 to 49 years (5). Despite low-income countries bearing a considerable burden, an increasing number of new syphilis infections have been observed in developed and emerging economies across North America, Europe, and the Asia-Pacific region (4).

Given the asymptomatic nature of syphilis, the need for early and accurate diagnostic methods becomes evident. Clinical manifestations of syphilis exhibit high variability depending on the disease stage, requiring careful interpretation of laboratory results within an individual’s medical history for accurate clinical diagnosis and effective case management (4). Laboratory diagnosis of syphilis involves either direct detection of *T. pallidum* or serological diagnosis of the infection (6). Direct detection methods, like dark-field microscopy and nucleic acid amplification testing, are useful in early primary syphilis but depend on the presence of clinical samples, which may not be available during a clinical visit. Moreover, these methods require highly trained laboratory technologists and sophisticated instruments, leading to variability in test performance (4, 6, 7). Therefore, serological testing remains the primary method for diagnosing syphilis infection due to the convenience of blood collection and the availability and affordability of serological assays. Serology tests for syphilis can be classified into (i) non-treponemal antibody tests such rapid plasma reagin [RPR] or Venereal Disease Research Laboratory test [VDRL]), which both detect Abs react with heart diphosphatidyl glycerol antigen (such as cardiolipin) produce by patients only during active TP infection and (ii) treponemal antibody tests detect antibodies react with specific TPA antigen. These antibodies can stay positive for life even after patient treatment; thus, they do not differentiate between active and past infection. Methods for detection of these antibodies includes TP particle agglutination (TPPA), TP hemagglutination assay (TPHA) fluorescent TP antibody absorption assay (FTA-ABS), ELISA, chemiluminescence immunoassay (CLIA). Figure 1 highlights the progression of treponemal and nontreponemal antibodies throughout the disease. A conclusive diagnosis of active (infectious) syphilis infection requires a combination of positive treponemal test (TT) and reactive non treponemal test (NTT) (8). However, each of the current TT and NTT methods has its limitations, hindering syphilis control and prevention efforts in real-world settings (4).

**Figure 1.**
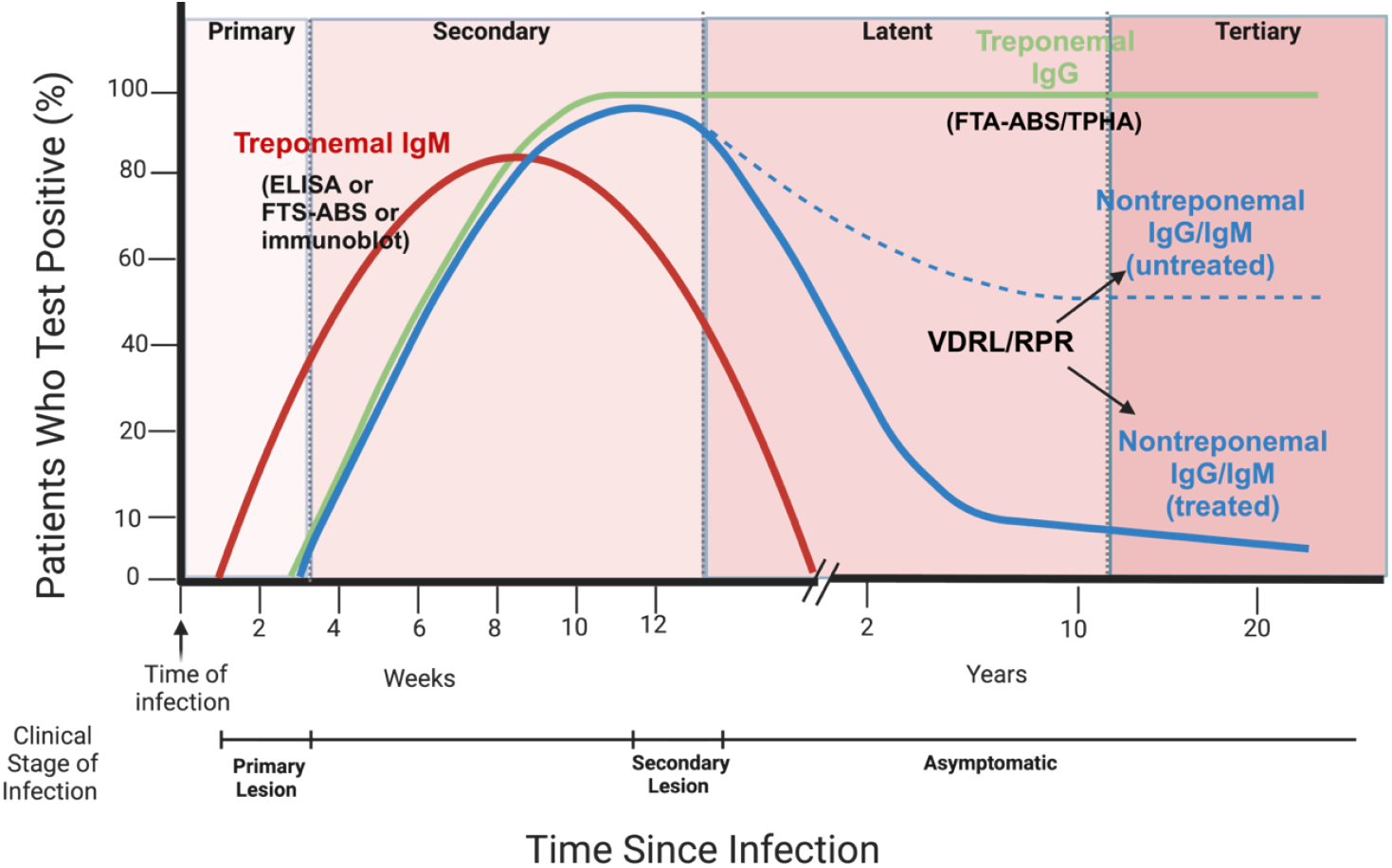
Progression of syphilis serology throughout the stages of the disease (adapted from (12)). This figure illustrates the serological response to Treponema pallidum infection, comparing treponemal and nontreponemal test results through disease stages. Treponemal tests detect IgG/IgM antibodies against T. pallidum antigens, becoming positive 2-4 weeks post-exposure and generally remaining reactive for life, indicating exposure but not necessarily active infection. Nontreponemal tests measure antibodies against lipoidal antigens from damaged cells/bacteria, reflecting disease activity and treatment efficacy as titers decrease post-therapy. Figure created by Biorender.

**Figure 2.**
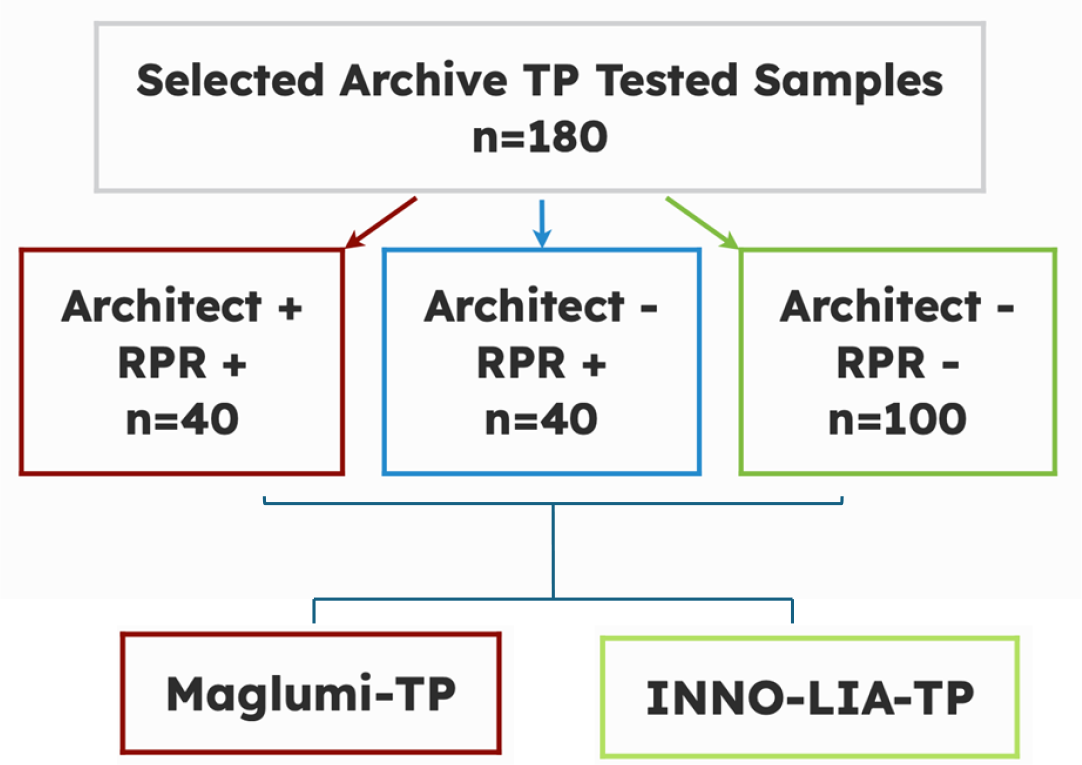
Flowchart depicting the sample selection process for inclusion in the study and evaluation approach.

There are two approaches for testing syphilis, which are (i) the traditional approach typically involves initial screening with a non-treponemal test, followed by confirmation with a treponemal-specific test and (ii) the reverse algorithm starts with a treponemal-specific test, followed by a non-treponemal test if the treponemal test is positive (9). Due to the advancement and the automation in ELISA and CLIA, the later approach has gained popularity because of its higher sensitivity and specificity.

In Qatar, infectious diseases are a major public health concern, accounting for approximately 8% of all deaths and significantly impacting the quality of life of its residents (9). The Qatar Ministry of Public Health (MOPH) plays a critical role in addressing these concerns through its Medical Commission (MC), which is tasked with the screening of newcomers to prevent the entry and spread of serious infectious diseases, including syphilis (caused by Treponema pallidum, TP). By law, all individuals entering the country must undergo a mandatory medical exam. The current diagnostic protocol implemented by the MC for the diagnosis of syphilis follows a traditional algorithm, beginning with a non-treponemal RPR test to screen for syphilis, followed by a confirmatory treponemal-specific test, typically using the Architect-TP assay, to identify active infections.

This study aims to critically assess the performance of the MAGLUMI®-X3 Syphilis assay (Maglumi-TP), as a potential sensitive and specific method for TP Ab detection. The validation was performed by comparing the Maglumi-TP results to the well-established and widely used ARCHITECT Syphilis TP (Architect-TP) CLIA treponemal assay and Fujiribio INNO-LIA® Syphilis Score (INNO-LIA-TP) line immunoassay. Both assays were utilized in our study as reference methods (10, 11).

## 2. Method

### 2.1 Ethical Approval

Our study was approved by the Institutional Review Board (IRB) at Qatar University (QU-IRB 017/2024-E).

### 2.2. Study Population and Study Design

We utilized samples previously collected by the Medical Commission (MC) in Qatar, operating under the MOPH, as part of routine screening for individuals entering the country. No patients or applicants were actively recruited for this study, and there was no direct or indirect interaction with any human subjects. In addition, to protect the participant confidentiality, we generated and assigned unique codes to each sample, categorizing them based on their testing outcomes to avoid the use of actual specimen barcode numbers that could potentially disclose the identities of the applicants. For example, samples were coded as N001 for those negative by both Architect-TP and RPR-TP, PN401 for samples positive by Architect-TP but negative by RPR-TP, and PP401 for samples positive in both the Architect-TP and RPR-TP assays.

We carefully selected 180 samples showing agreement and discrepancies between RPR and Architect-TP methods. This selection included 40 cases positive by both methods, 40 cases positive by RPR but negative by Architect-TP, and 100 cases negative by both assays. All selected samples underwent re-testing using the Maglumi-TP assay and were further confirmed by the INNO-LIA-TP assay.

### 2.3. Architect-TP assay

The Architect-TP assay, developed by Abbott Diagnostics in Abbott Park, Illinois, USA, is a two-step immunoassay designed for the qualitative detection of antibodies to *T. pallidum* in human serum or plasma. This assay employs chemiluminescent microparticle immunoassay (CMIA) technology with adaptable assay protocols known as Chemiflex (13). The assay was performed following the manufacture protocol (13). Briefly, 80 µL of the sample is mixed with microparticles coated with recombinant TP antigens (TpN15, TpN17, and TpN47). Anti-TP antibodies in the sample bind to the TP-coated microparticles, then, acridinium-labeled anti-human IgG and IgM conjugate are introduced to create a reaction mixture. The resulting chemiluminescence is measured in RLUs, with nonreactive samples exhibiting S/CO values less than 1.00, and reactive samples having S/CO values equal to or greater than 1.00.

### 2.4. Maglumi-TP assay

The Maglumi-TP assay (Snibe Diagnostics Co. Ltd., Shenzhen, China) is a sandwich CLIA. Briefly, 80 µL of the sample was added to ABEI labeled with *T. pallidum*-specific recombinant antigens (TpN47, TpN17, and TpN15), along with magnetic microbeads coated with *T. pallidum* -specific recombinant antigens. The total *T. pallidum* antibodies in the sample form a sandwich complex with the recombinant antigens. The resulting light signal, measured as RLUs, is proportional to the concentration of total antibodies against TP. A result greater than or equal to 1.0 mIU/mL (≥1.0 mIU/mL) is considered to be positive.

### 2.5. INNO-LIA-TP assay

INNO-LIA-TP (Fujirebio Europe N.V., Ghent, Belgium), a LIA for the simultaneous detection of antitreponemal antibodies, makes use of recombinant antigens and synthetic peptide antigens derived from *T. pallidum*. Three recombinant antigens (TpN47, TpN17, and TpN15) and one synthetic peptide (TmpA) serve as antigens in this assay. The assay was conducted following the manufacturer’s protocol. In brief, serum or plasma samples were diluted at a ratio of 1:100 and incubated at room temperature (20°C) overnight. This incubation was followed by three washing steps before the addition of a goat anti-human IgG conjugated to alkaline phosphatase. After another round of incubation, three washing steps were performed, followed by the addition of a chromogen.

After color development, each line was compared to the control lines, and the intensities were scored as follows: 0: No line or a line less intense than the cutoff line, 1: A line with an intensity between that of the cutoff line and the 1+ control line, 2: A line with an intensity between that of the 1+ control line and the 3+ control line, 3: A line equal to that of the 3+ control line, 4: A line with an intensity greater than that of the 3+ control line

A sample is considered TP antibody negative if no band or a single band with a maximum intensity of 0.5 is present. If multiple bands with a minimum intensity of 0.5 are visible, the sample is considered TP antibody positive. Finally, a sample is considered indeterminate if a single band is visible with a minimum intensity of 1. The results were captured and interpreted using the LiRAS software designed for result interpretation in Line Immunoassays for infectious diseases.

#### Rapid Plasma Reagin test (RPR)

The RPR card test, developed by Fortress Diagnostics Limited, Antrim, UK, is a qualitative and semi-quantitative non-treponemal flocculation assay designed to detect reagin antibodies in patient serum. The assay was conducted following the manufacturer’s protocol (14). In brief, 50 μL of the patient’s serum or plasma was dispensed and spread to cover a defined circle on the RPR test card. Subsequently, add one drop of the antigen (provided by the manufacturer) to the sample and mix by rotation at 100 rpm for 8 minutes on an automatic rotator. Qualitative interpretation included reactive results with medium and large aggregates, weakly reactive with finely dispersed aggregates, and non-reactive with no visible aggregates and only a smooth grey appearance. Positive samples underwent semi-quantitative analysis, involving serial dilutions (50 μL) of the patient’s serum with isotonic saline.

### 2.7. Statistical analysis

Descriptive statistical analysis was conducted for categorical variables. Concordance analysis between Maglumi-TP assay and the other assays was performed, including sensitivity, specificity, overall percent agreement (OPA), positive percent agreement (PPA), negative percent agreement (NPA), positive predictive value (PPV), negative predictive value (NPV), and Cohen’s Kappa test. Interpretation of Cohen’s Kappa coefficient values was as follows: k ≤ 0 indicating no agreement, 0.01–0.20 representing none to slight agreement, 0.21–0.40 denoting fair agreement, 0.41–0.60 indicating moderate agreement, 0.61–0.80 reflecting substantial agreement, and 0.81–1.00 signifying almost perfect agreement (15, 16). GraphPad Prism (Version 9, San Diego, CA, USA) was utilized for the execution of all statistical tests.

## 3. Results

### 3.1. Architect-TP assay showed perfect concordance with INNO-LIA-TP

A comparative analysis was conducted between the well-established treponemal assay CLIA Architect-TP assay and the INNO-LIA-TP line immunoassay as a reference assay. Out of the 180 samples examined, 39 (21.7%) were identified as true positives, and 139 (77.2%) were confirmed as true negatives as shown in Table 1. Notably, the Architect-TP assay demonstrated exceptional performance, with no instances of false negatives or false positives compared to the INNO-LIA-TP.

**Table 1.**
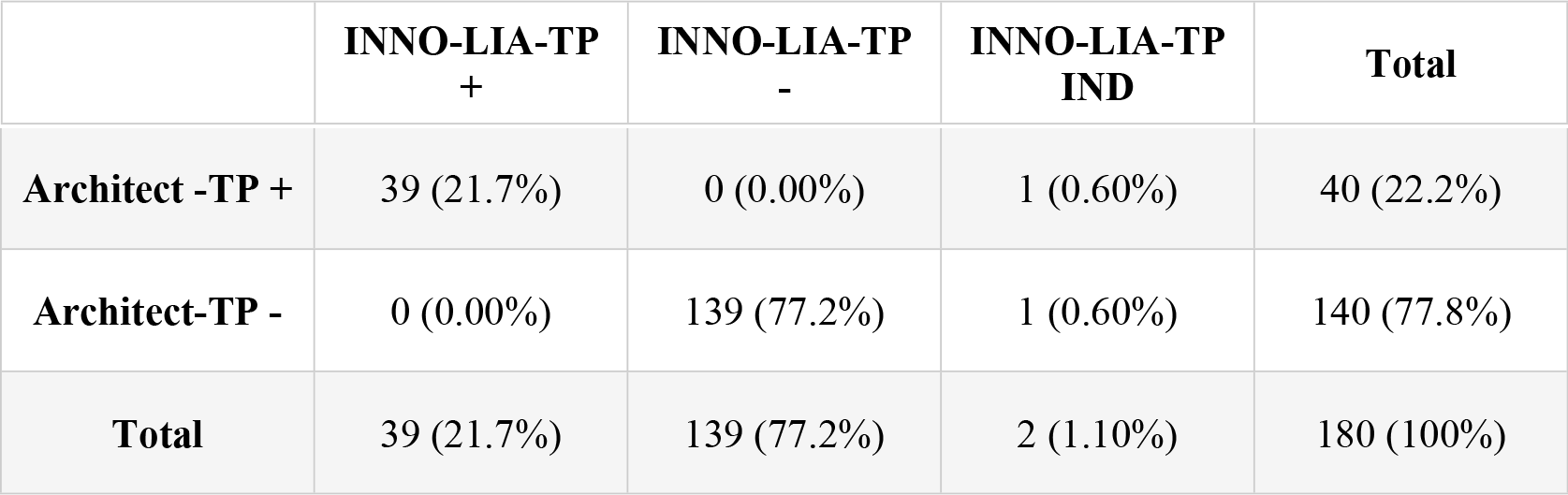
Comparison of Architect-TP results with INNO-LIA-TP (n=180)

|  | INNO-LIA-TP<br>+ | INNO-LIA-TP<br>- | INNO-LIA-TP<br>IND | Total |
| --- | --- | --- | --- | --- |
| **Architect -TP +** | 39 (21.7%) | 0 (0.00%) | 1 (0.60%) | 40 (22.2%) |
| **Architect-TP -** | 0 (0.00%) | 139 (77.2%) | 1 (0.60%) | 140 (77.8%) |
| **Total** | 39 (21.7%) | 139 (77.2%) | 2 (1.10%) | 180 (100%) |

### 3.2. Maglumi-TP assay demonstrates perfect concordance with INNO-LIA-TP

A comprehensive analysis was conducted to compare the Maglumi-TP assay with INNO-LIA-TP. Among the 180 samples, 39 (21.7%) were identified as true positives, and 138 (76.7%) were confirmed as true negatives. The Maglumi-TP assay exhibited exceptional performance, showing perfect concordance with INNO-LIA-TP, with no occurrences of false negatives or false positives (Table 2).

**Table 2.**
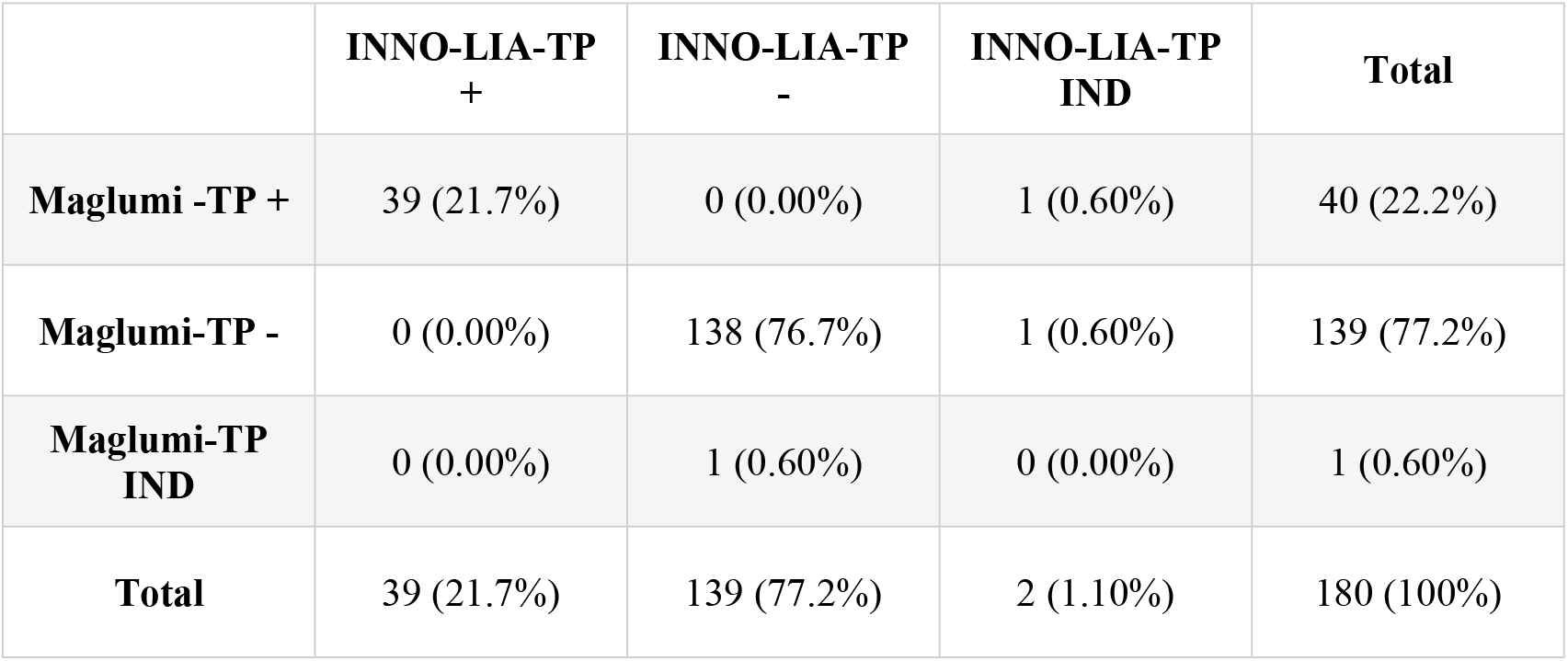
Comparison of Maglumi-TP results with INNO-LIA-TP (n=180)

### 3.4. Maglumi-TP assay Demonstrates Perfect Concordance with Architect-TP

A comparative analysis was conducted between Maglumi-TP assay and Architect-TP. Among the 180 samples tested 40 (22.2%) were identified as true positives, and 139 (77.2%) were confirmed as true negatives as shown in Table 4. The Maglumi-TP assay exhibited exceptional performance, demonstrating perfect concordance with the Architect-TP, with no instances of false-negative or false-positive results.

**Table 4.** Comparison of Maglumi-TP results with Architect-TP (n=180)

|  | **Architect-TP +** | **Architect-TP-** | **Total** |
| --- | --- | --- | --- |
| **Maglumi -TP +** | 40 (22.2%) | 0 (0.00%) | 40 (22.2%) |
| **Maglumi-TP -** | 0 (0.00%) | 139 (77.2%) | 139 (77.2%) |
| **Maglumi-TP IND** | 0 (0.00%) | 1 (0.60%) | 1 (0.60%) |
| **Total** | 40 (22.2%) | 140 (77.8%) | 180 (100%) |

### 3.5. Maglumi-TP assay showed excellent performance similar to the well-established Architect-TP assay in comparison to the INNO-LIA-TP

In the comprehensive evaluation of Maglumi-TP and Architect-TP against the reference assay INNO-LIA-TP, both assays demonstrated outstanding performance. High sensitivity, specificity, OPA, PPV, and NPV were observed for both assays, each achieving 100.0% with narrow confidence intervals (95% CI: 91.0-100.0%, 97.4-100.0%, 97.9-100.0%, 91.0-100.0%, and 97.4-100.0%, respectively) as depicted in Table 5. The Cohen’s kappa statistic (*κ* = 1.00) indicated excellent agreement between Maglumi-TP and INNO-LIA-TP.

**Table 5.** Performance Evaluation of commercially available assays in comparison to the reference method; INNO-LIA-TP *.

| **Referen**<br>**ce** | **Test** | **Sensitiv**<br>**y %** | **Specificit**<br>**y %** | **PPV%** | **NPV%** | **PPA%** | **NPA%** | **OPA%** | **Cohen's**<br>**Kappa**<br>**Coefficie**<br>**nt %n** |
| --- | --- | --- | --- | --- | --- | --- | --- | --- | --- |
| **INNO-LIA-TP** | **Architect-TP** | 100 (91-100) | 100 (97.4-100) | 100 (91-100) | 100 (97.4-100) | 100 (91-100) | 100 (97.4-100) | 100 (97.9-100) | 1.00 (0.00-1.00) |
| **INNO-LIA-TP** | **Maglumi-TP** | 100 (91-100) | 100 (97.4-100) | 100 (91-100) | 100 (97.4-100) | 100 (91-100) | 100 (97.4-100) | 100 (97.9-100) | 1.00 (0.00-1.00) |
| **Architect-TP** | **Maglumi-TP** | 100 (91.2-100) | 100 (97.4-100) | 100 (91.2-100) | 100 (97.4-100) | 100 (91.2-100) | 100 (97.4-100) | 100 (98-100) | 1.00 (0.00-1.00) |

*\*Samples with indeterminate results were excluded from the evaluation.*

**Table 6.**
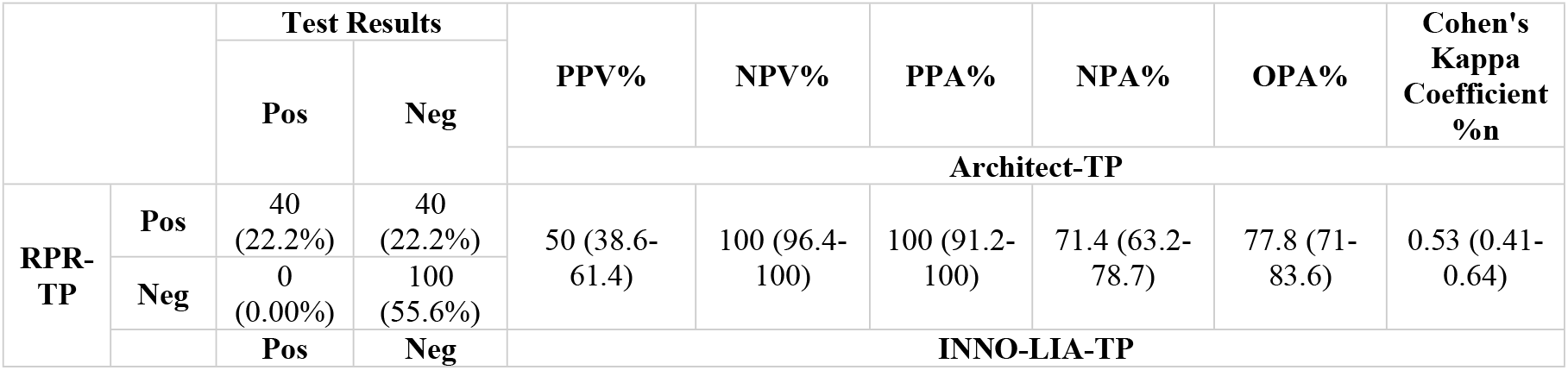
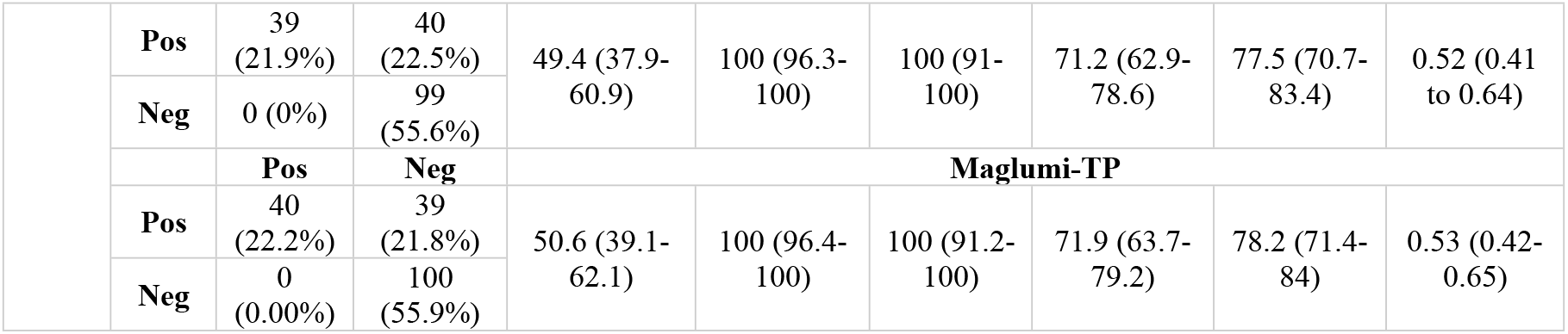
Comparison RPR results with INNO-LIA-TP, Architect-TP, and Maglumi-TP (n=180).

### 3.3. RPR assay demonstrates high false positive rate compared to the INNO-LIA-TP, Architect-TP, and Maglumi-TP

A comprehensive comparative analysis was conducted to evaluate the performance of the RPR assay compared to INNO-LIA-TP, which served as the reference method. Among the 180 samples evaluated, 39 (21.7%) were confirmed as true positives, while 99 (55.0%) were verified as true negatives as shown in Table 3. Notably, 40 (28.8%; 40/139) exhibited false-positive results when using the RPR test compared to the INNO-LIA-TP, indicating a substantial false positive rate associated with the RPR assay.

In the comprehensive assessment of RPR against the reference assay INNO-LIA-TP, RPR exhibited weak/poor performance. The OPA, PPV, and NPV were calculated as 77.5% (95% CI: 70.7-83.4), 49.4% (95% CI: 37.9-60.9), and 100% (95% CI: 96.3-100) respectively. The Cohen’s kappa statistic (*κ* = 0.50) indicated fair agreement between RPR and INNO-LIA-TP.

## 4. Discussion

Throughout the 20th century, syphilis serology testing played a crucial role in limiting the spread of the venereal disease (17). The reactivity of a non-treponemal test, like RPR, holds critical significance as an indicator of disease activity, particularly from a public health standpoint. On the other hand, the reactivity of a treponemal test cannot distinguish between current and past infections. Nevertheless, RPR plays a vital role in monitoring treatment efficacy and assessing disease recurrence, making both tests indispensable for accurate syphilis diagnosis. Traditionally, laboratory diagnosis started with a nontreponemal test, such as RPR, followed by a Treponema pallidum–specific assay, like T. pallidum particle agglutination assay (TP-PA). Recently, automated treponemal-specific immunoassays (e.g., EIAs, CIAs, and microbead immunoassays) have gained prominence due to their efficiency in reducing labor and turnaround time (18). Because modern treponemal immunoassays have demonstrated equivalent sensitivity and specificity to nontreponemal assays (apart from early primary syphilis) combined with their ability to be automated using high-throughput instrumentation, many laboratories have implemented the reverse algorithm, beginning with a treponemal immunoassay, followed by nontreponemal testing on initially reactive specimens (19). The advantages of the reverse algorithm can include detection of early infection (before nontreponemal antibodies can be detected), latent infection (after nontreponemal antibodies have disappeared), automated workflow, and objective results reporting (6). Currently, the Centers for Disease Control and Prevention (CDC) recommends conducting a TP-PA only if there are discordant results between the immunoassay and RPR (eg, EIA-reactive, RPR-nonreactive) (19). Regardless of the chosen algorithm, it is essential for laboratories to consider the sensitivity and specificity of treponemal tests when selecting the most appropriate approach.

The MC laboratory adheres to the traditional syphilis screening algorithm, beginning with RPR as the primary screening approach, followed by a treponemal test (ARCHITECT® Syphilis TPA assay) as a confirmatory assay for reactive samples (20, 21). Our previous study raised concerns about relying solely on RPR results due to a high false-positive rate (22). The PPV of the RPR test was found to be only 36.5%, with over 50% of RPR-reactive samples being negative in the TPA confirmatory test (22). Similarly, in the current study, RPR demonstrated low PPV of 44.9% and a substantial false-positive rate of 28.8% compared to INNO-LIA-TP, emphasizing the limitations of RPR, particularly in terms of specificity (71.2%). The fair overall agreement (Cohen’s kappa of 0.50) underscores the importance of cautious interpretation of RPR results for syphilis diagnosis. RPR tests may show false positive results for various reasons, including lupus, viral mononucleosis, malaria, leprosy, viral pneumonia, and rickettsia infection.

The widely employed Abbott Architect-TP, known for its automation capabilities, increased throughput, and high sensitivity and specificity (23-27), demonstrated remarkable performance in our study, achieving 100% sensitivity and specificity when INNO-LIA-TP served as the reference test. Aligning with our findings, a multicenter study conducted across 13 independent laboratories in 10 diverse regions in China reported that the Architect-TP demonstrated a sensitivity of 98.26% and specificity of 99.74% (28). Another study affirmed the Architect-TP’s high performance, with sensitivity and specificity reaching 98.80% and 99.58%, respectively (24). Additionally, Tao C et al reported a PPV of 88.25% and a NPV of 99.97% (28). Interestingly, our study revealed superior PPV and NPV values, suggesting potential variations based on the studied population.

The Maglumi-TP immunoassay exhibited comparable specificity and sensitivity, reaching 100% for routine sample screening, similar to the widely utilized Architect-TP. Remarkably, Maglumi demonstrates neither false negatives nor false positives. In our practical experience, the user-friendly and reliable Maglumi X3 CLIA system, with a throughput of 72 tests per hour and a testing duration of just 18 minutes, proves to be a time-effective and convenient option for syphilis diagnostics in high-volume clinical laboratories.

Among the three treponemal assays, discrepancies were observed in three samples. Firstly, one sample exhibited an indeterminate result in INNO-LIA-TP, with a positive TpN17, while both Maglumi-TP and Architect yielded positive results. This could be indicative of the high sensitivity of the TpN17 antigen in detecting primary and secondary syphilis, as documented by Rostopira et al., who reported 100% sensitivity of TpN17 in these stages (29). The indeterminate result in INNO-LIA-TP might suggest early or borderline reactivity, where TpN17’s high sensitivity could detect syphilis earlier than other antigens. In the second case, the sample tested negative by both Architect-TP and INNO-LIA-TP but exhibited an indeterminate result with Maglumi-TP (Concentration: 1.3 mIU/mL), likely representing a false positive result. Notably, this sample also demonstrated a positive result in the RPR test. Lastly, the third sample returned negative results in all treponemal assays and RPR, except for an indeterminate result with INNO-LIA-TP linked to the TmpA antigen, likely indicating false positive result. Previous studies have shown that certain antigens like TmpA can cross-react with sera from patients with conditions such as mononucleosis, hepatitis, diabetes mellitus, HIV/AIDS, and old age, which could explain the discrepancies observed in the second and third cases (30).

One potential limitation of our study is the relatively small sample size, and the subjectivity associated with sample selection based on Architect-TP and RPR results. Furthermore, our study did not investigate other potential factors that might influence assay performance, such as variations in sample storage conditions or the presence of interfering substances or the stage of the disease. Addressing these factors in future research endeavors, beside evaluating the current screening protocol of using traditional algorithm against the Reverse algorithm in screening setting like medical commission dept, would contribute to a more comprehensive understanding of assay dynamics and refine the interpretation of diagnostic outcomes and the impact on public health.

## 5. Conclusion

Our study provides a comprehensive performance evaluation of Maglumi-TP for syphilis detection in comparison to the current screening method, Architect-TP. The results demonstrate that Maglumi-TP exhibits excellent sensitivity and specificity, comparable to the widely established Architect-TP. Furthermore, Maglumi-TP shows strong agreement with INNO-LIA-TP, indicating its reliability in confirming syphilis infection. These findings highlight the robustness and accuracy of Maglumi-TP in detecting syphilis and support its adoption as a reliable diagnostic tool. For routine screening of blood samples, it offers the advantage of being fast, easy to use, and suitable for high-throughput platforms. However, like other treponemal assays, the Maglumi-TP is most suitable for the diagnosis of recently acquired and previously treated infections. Finally, our findings also affirm the robust performance of Architect-TP within our existing testing protocol, evidenced by its excellent sensitivity, specificity, and consistent agreement with INNO-LIA-TP.

## Data Availability

All relevant data are within the manuscript and its Supporting Information files.

## 6. Acknowledgement

All immunoassays used were provided from the corresponding companies to GKN lab as in-kind support (free of charge) for this study. Additionally, we would like to acknowledge the technicians in the MC for conducting all testing.

Conceptualization: AI; Methodology: N.Y., J.A.A., H.M.K., M.N.A., M.E., I.W.K., M.A.I., M.M.A., P.B.N., S.Y.,; Formal Analysis: G.K.N., N.Y., L.J.A.-R., A.I; Validation: G.K.N., L.J.A.-R., AI.,; Investigation: G.K.N., H.M.Y., L.J.A.-R., A.I; Resources: G.K.N., A.I.; Data Curation: G.K.N., N.Y., L.J.A.-R., A.I.; Writing—Original Draft Preparation: G.K.N., N.Y.,; Writing— Review and Editing: G.K.N., N.Y., H.M.Y., M.N.A., M.E., I.W.K., M.A.I., M.M.A., I.A.S., S.Y. L.J.A.-R., A.I.; Visualization: G.K.N., N.Y., L.J.A.-R., AI..,; Supervision: A.I.,; Project Administration: A.I., Funding Acquisition: G.K.N., L.J.A.-R. All authors have read and agreed to the published version of the manuscript.

## Ethics approval statement

The study was reviewed and approved by the Institutional Review Board at Qatar University (QU-IRB 017/2024-E).

